# Comparative analysis of functional assay evidence use by ClinGen Variant Curation Expert Panels

**DOI:** 10.1101/19000661

**Authors:** Dona M. Kanavy, Shannon M. McNulty, Meera K. Jairath, Sarah E. Brnich, Chris Bizon, Bradford C. Powell, Jonathan S. Berg

## Abstract

**Background:** The 2015 American College of Medical Genetics and Genomics (ACMG) and Association for Molecular Pathology (AMP) guidelines for clinical sequence variant interpretation state that “well-established” functional studies can be used as evidence in variant classification. These guidelines articulated key attributes of functional data, including that assays should reflect the biological environment and be analytically sound; however, details of how to evaluate these attributes were left to expert judgment. The Clinical Genome Resource (ClinGen) designates Variant Curation Expert Panels (VCEPs) in specific disease areas to make gene-centric specifications to the ACMG/AMP guidelines, including more specific definitions of appropriate functional assays. We set out to evaluate the existing VCEP guidelines for functional assays.

**Methods:** We evaluated the functional criteria (PS3/BS3) of six VCEPs (*CDH1*, Hearing Loss, Inherited Cardiomyopathy-*MYH7, PAH, PTEN*, RASopathy). We then established criteria for evaluating functional studies based on disease mechanism, general class of assay, and the characteristics of specific assay instances described in primary literature. Using these criteria, we extensively curated assay instances cited by each VCEP in their pilot variant classification to analyze VCEP recommendations and their use in the interpretation of functional studies.

**Results:** Unsurprisingly, our analysis highlighted the breadth of VCEP-approved assays, reflecting the diversity of disease mechanisms among VCEPs. We also noted substantial variability between VCEPs in the method used to select these assays and in the approach used to specify strength modifications, as well as differences in suggested validation parameters. Importantly, we observed discrepancies between the parameters VCEPs specified as required for approved assay instances and the fulfillment of these requirements in the individual assays cited in pilot variant interpretation.

**Conclusions:** Interpretation of the intricacies of functional assays often requires expert-level knowledge of the gene and disease and current VCEP recommendations for functional assay evidence are a useful tool to improve the accessibility of functional data. However, our analysis suggests that further guidance is needed to standardize this process and ensure consistency in the application of functional evidence.

## BACKGROUND

In 2015, the American College of Medical Genetics and Genomics (ACMG) and Association for Molecular Pathology (AMP) established standards and guidelines [1] for clinical variant interpretation. These guidelines provided criteria for classifying variants as pathogenic (P), likely pathogenic (LP), variant of uncertain significance (VUS), likely benign (LB), or benign (B) using distinct evidence types, each of which was assigned a level of strength. Additional rules specified combinations of the types and strengths of criteria sufficient to reach a pathogenic or benign classification. In cases of insufficient or conflicting evidence, variants were classified as VUS, which present a challenge in clinical molecular genetic testing as they should not alone be used to define clinical decision-making according to ACMG/AMP standards. Functional data has considerable potential to aid in variant classification, particularly VUS reclassification [2]. In contrast to the opportunistic nature of many types of evidence (such as the fortuitous discovery of a family with sufficient segregation data to aid interpretation), functional assays are the most amenable to development and therefore the most tractable to be produced in a timely manner after a variant is observed. The ACMG/AMP guidelines state that results of “well-established” functional studies can qualify as evidence for functional criteria application coded as PS3 or BS3 (an abbreviation for functional evidence in the direction of a pathogenic or benign interpretation, respectively, at a default evidence strength of strong) and that validation, reproducibility, robustness, and ability of the assay to reflect the biological environment should be considered. However, it is unclear how these attributes should be evaluated and selecting appropriate functional evidence often requires expert-level knowledge of the gene and disease.

The Clinical Genome Resource (ClinGen) has founded Variant Curation Expert Panels (VCEPs) in multiple disease areas, each tasked with developing adaptations of the ACMG/AMP rules for their disease or gene of interest [3]. These VCEP specifications regarding functional data provided expert interpretations of the qualities required for an assay to be deemed “well-established.”

In this study, we sought to define the characteristics of functional assays that satisfy PS3/BS3 criteria by conducting a comparative analysis of VCEP recommendations for these criteria. The six VCEPs that have published disease- and gene-specific adaptations to the ACMG/AMP guidelines (*CDH1*, Hearing Loss, Inherited Cardiomyopathy-*MYH7, PAH, PTEN*, and RASopathy [3–8]) served as a case study, allowing us to assess the validation parameters and evidence strength for each approved assay, as well as the features of assays that were not approved by the VCEPs. We curated instances of assays in the primary literature cited by each VCEP both in their recommendation publication and in the course of their pilot variant classification using consistent criteria. This approach allowed us to assess the extent to which cited instances of assays satisfy VCEP-specified recommendations and how they differed.

## METHODS

### Evaluation of ClinGen VCEP Specifications

We assessed the guidance for the use of PS3/BS3 by six ClinGen VCEPs with approved and published variant interpretation recommendations as of April 2019: *CDH1*, Hearing Loss, Inherited Cardiomyopathy (*MYH7*), *PAH, PTEN*, and RASopathy. In our initial survey of recommendations, we noticed certain parameters (replicates, controls, thresholds, and validation measures) were identified by more than one group. We evaluated how often these four assay parameters were specified by the VCEPs and whether each VCEP provided recommendations for modifying PS3/BS3 evidence strength to a moderate (PS3_M) or supporting (PS3_P/BS3_P) level.

### Literature Search and Variant Identification

To identify relevant primary literature for each VCEP, we catalogued each of the variants classified by the VCEP as part of their pilot variant classification effort and the final classification of each pilot variant (P, LP, VUS, LB, or B). Next, we determined which pilot variant interpretations included PS3/BS3 evidence and the specific instances of assay cited as evidence utilizing information in the VCEP publication, as well as ClinVar (https://www.ncbi.nlm.nih.gov/clinvar/) and the ClinGen Evidence Repository (https://erepo.clinicalgenome.org/evrepo/). In addition to curating primary literature cited as evidence in pilot variant interpretation, we curated primary literature and reviews the VCEPs cited in their publications in support of their approval or exclusion of a given assay (see Literature Curation Approach).

### Inclusion and Exclusion Criteria

We focused our curation efforts on assays that determined the function of a gene product. We excluded assays that tested splicing, as these typically evaluated the transcript rather than the encoded protein function. We conducted a limited evaluation of assay instances using cells or tissue derived from affected individuals as the primary experimental material, as the variant in question was not isolated from the individual’s genetic background and, as a result, abnormal gene product function cannot be definitively attributed to the genetic variant.

### Literature Curation Approach

We developed consistent criteria for evaluating classes of functional assays and specific instances of their use in evaluating the impact of a variant by establishing three main domains to describe a given assay. First, we curated the disease mechanism for a given gene-disease pair using the associated Monarch Disease Ontology (MONDO) identifier [10], the functional pathway using Gene Ontology (GO) terms [11–13], the molecular etiology using controlled vocabulary (e.g., loss-of-function, dominant negative, or gain-of-function), and the inheritance pattern, also using a controlled vocabulary. Next, we identified the general class of each assay used in the primary literature each VCEP cited using ontology terms from Bioassay Ontology (BAO; http://bioassayontology.org/) [14,15] and Evidence and Conclusion Ontology (ECO; http://www.evidenceontology.org/) [16]. In some cases, ontologies describing the class of assay were found in only one of the two ontology databases. Finally, we used a structured narrative to describe the specific instance of an assay being performed. We summarized multiple attributes, including PubMed Identifier (PMID), study purpose, entity performing the assay, methodology (including replicates, controls, thresholds, and validation measures), and assay results. We also catalogued other details specific to the assay, such as experimental material, quantitation measures, and statistical analyses.

## RESULTS

Each VCEP approved between one and seven assays for use as evidence for PS3/BS3 application (Table 1), all reflective of the disease mechanism but with widely varying specificity regarding the descriptions of approved assays. These ranged from detailed assays evaluating the myristoylation status of a single residue in a given protein (RASopathy VCEP) to broader specification of any mammalian variant-specific knock-in model (Inherited Cardiomyopathy VCEP). Two VCEPs (Hearing Loss and *PTEN*) approved any sufficiently validated assays not explicitly approved in their recommendations if deemed appropriate by the analyst in future variant interpretation efforts. We also noted variability in the inclusion of guidance for downgrading strength modifications to a moderate or supporting level. We next surveyed the parameters stipulated by each VCEP (Table 2). We also observed variation in the frequency and methods by which these parameters were specified, with most VCEPs detailing a need for one to two of these four parameters to be fulfilled by an individual instance of a functional assay.

**Table 1:**
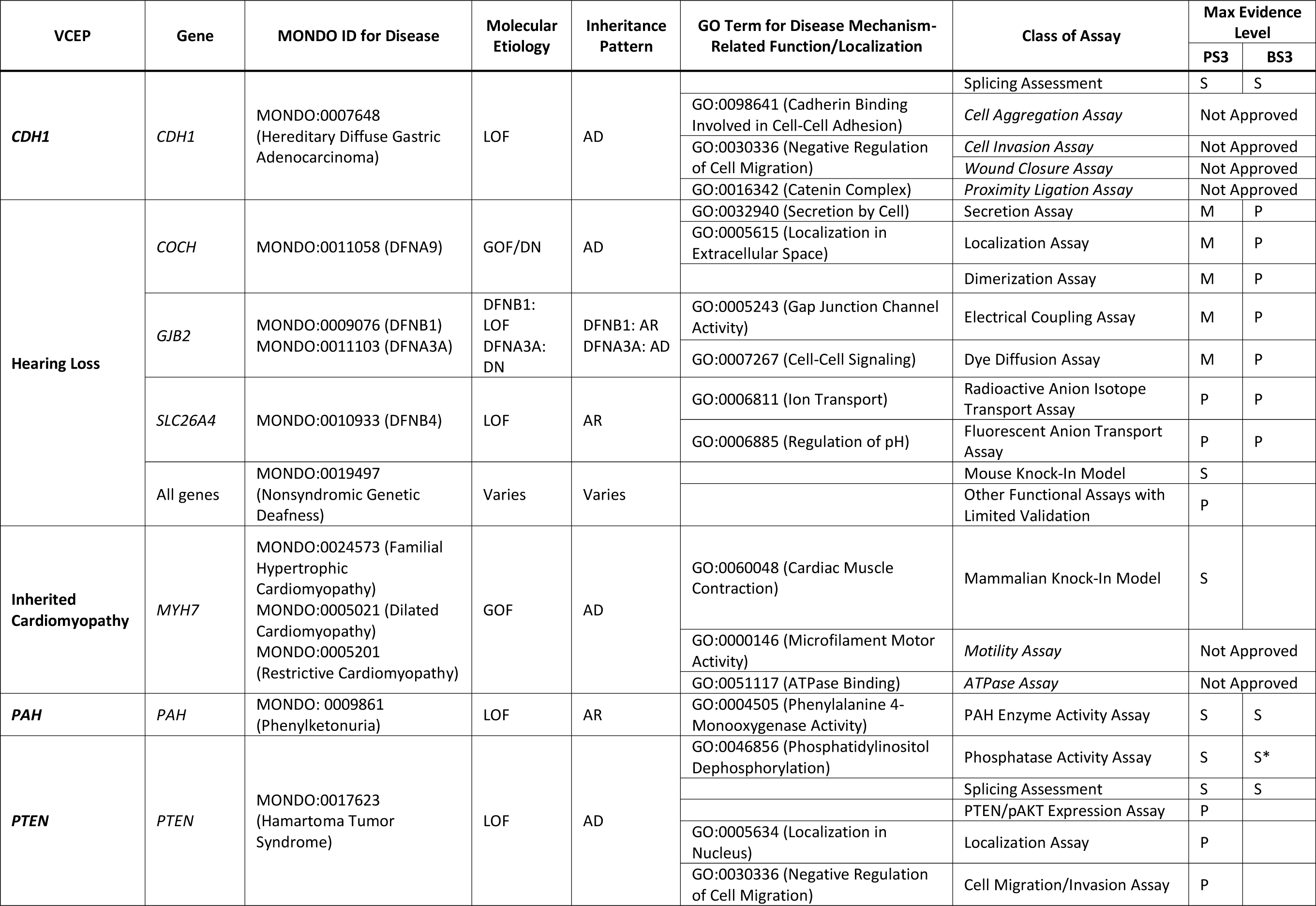
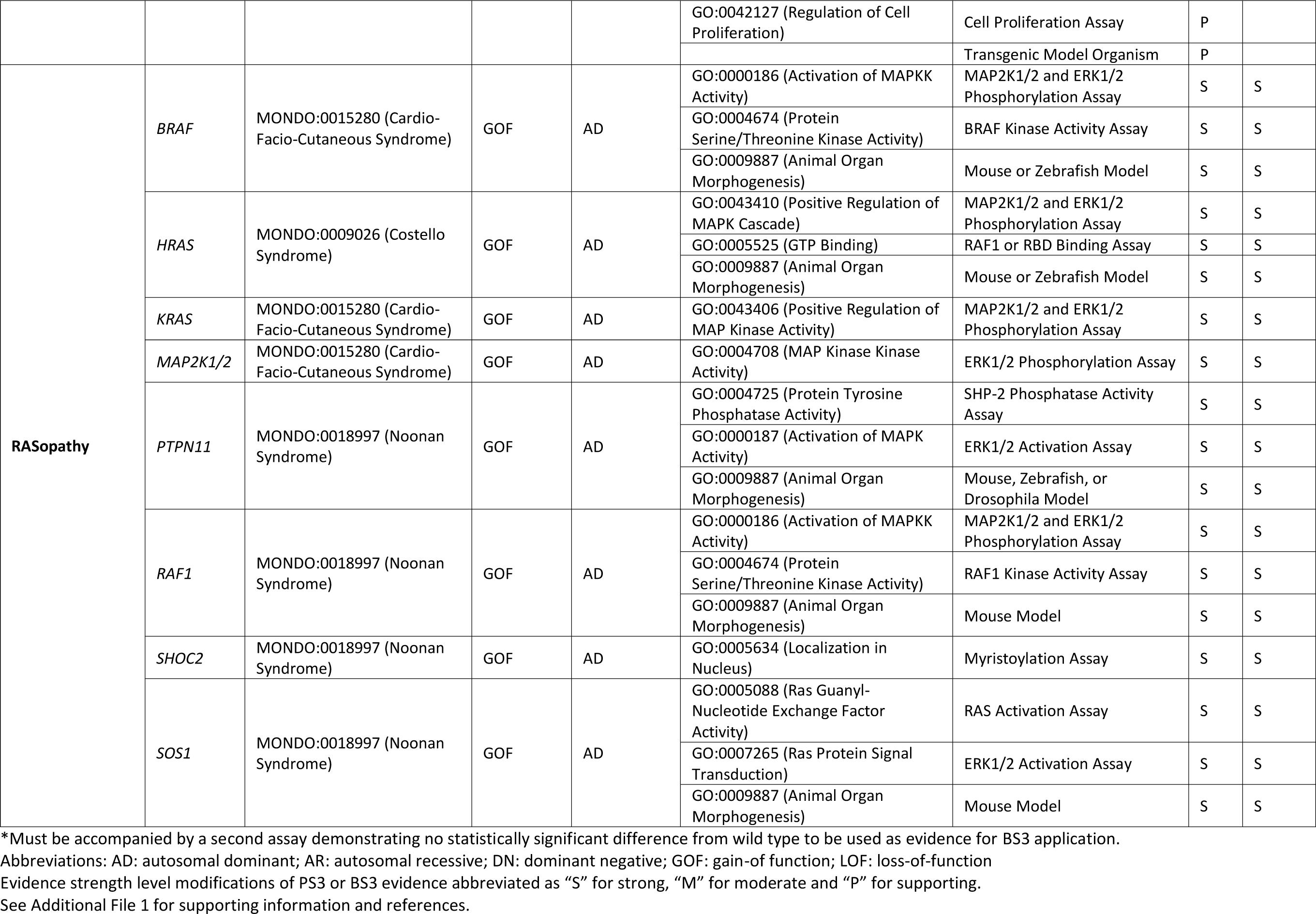
Overview of Variant Curation Expert Panel (VCEP) disease areas and mechanisms, functional assay classes, and accompanying strength level modifications.

**Table 2:** Summary of PS3/BS3 assay parameter specifications and strength modification recommendations by each Variant Curation Expert Panel (VCEP).

| VCEP | Replication | Controls | Thresholds | Validation | Possible PS3 Evidence Levels |
| --- | --- | --- | --- | --- | --- |
| ***CDH1*** |  |  | ✓ | ✓ | Strong, supporting |
| **Hearing Loss** |  | ✓ |  | ✓ | Moderate, supporting |
| **Inherited Cardiomyopathy (*MHY7*)** |  |  |  | ✓ | Strong |
| ***PAH*** | ✓ |  |  |  | Strong |
| ***PTEN*** | ✓ | ✓ | ✓ |  | Strong, supporting |
| **RASopathy** |  |  |  | ✓ | Strong |

The frequency of functional criteria application in pilot variant interpretation varied widely among VCEPs (Fig. 1a), with the *PAH* and RASopathy VCEPs using PS3/BS3 at the highest frequency in their pilot variant classification (31/85 variants and 36/103 variants, respectively) while the *CDH1* and Inherited Cardiomyopathy VCEPs applied PS3/BS3 less commonly (4/49 variants and 4/60 variants, respectively). Variants that were ultimately classified as VUS rarely included PS3/BS3 evidence codes (Fig. 1b). We noted general agreement between the functional data criteria applied to pilot variants and the overall variant classification (Fig. 1c). Pilot variant interpretations that included PS3 criteria were frequently given an overall classification of LP or P, with very few classified as VUS and none classified as LB or B. Similarly, those that included BS3 criteria were often classified as LB or B, with one interesting exception of a variant with BS3 evidence ultimately classified as P. Given the variation observed in our broad analysis of parameter specification across VCEPs, we used consistent criteria to curate primary literature cited by each of the six VCEPs to assess their application of these parameters (see Methods).

**Fig. 1.**
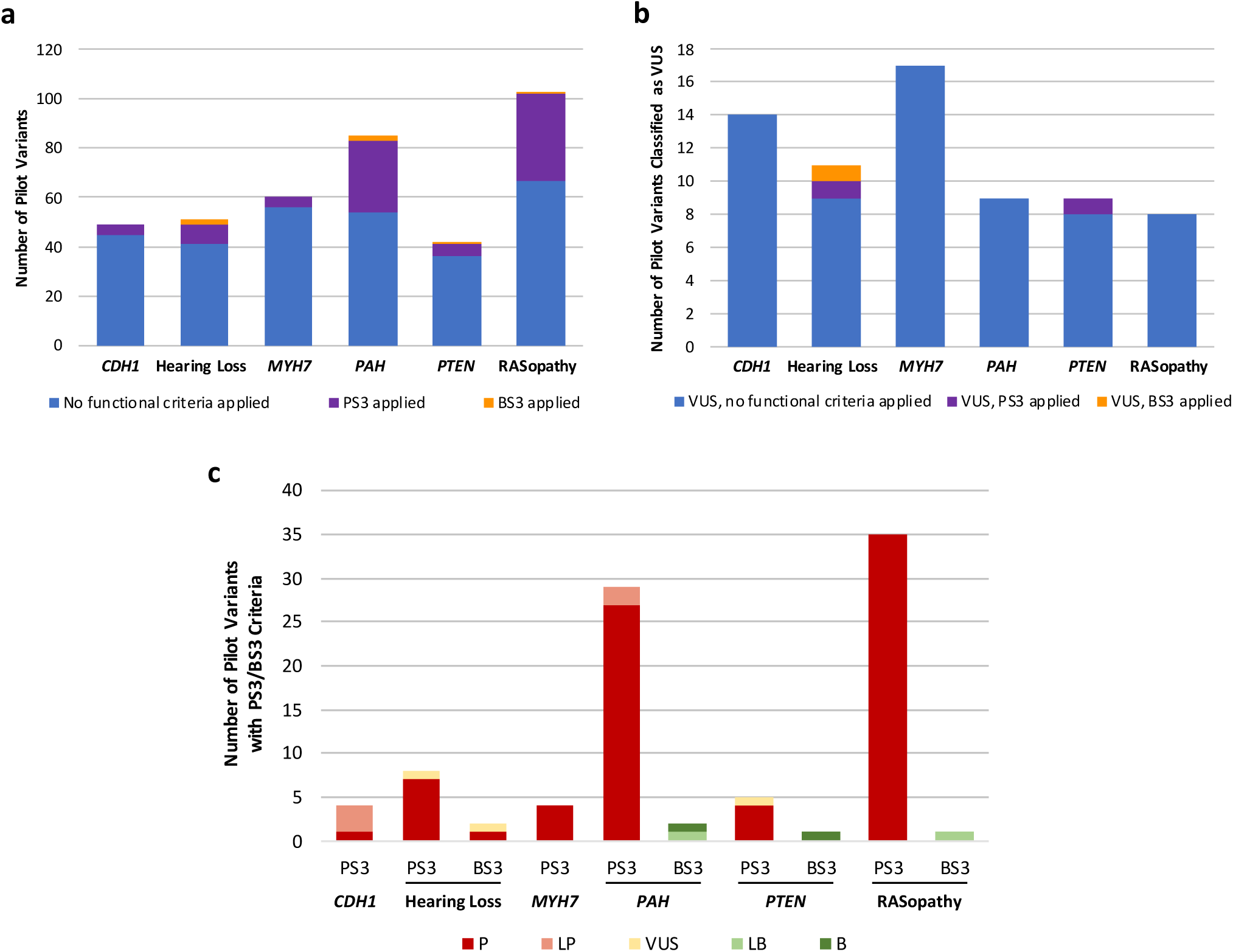
Use of the PS3/BS3 criteria in Variant Curation Expert Panel (VCEP) pilot variant classification. **a** Comparison of PS3 criterion application at any strength level (purple) and BS3 criterion application at any strength level (orange) in the pilot variant classification of each VCEP. **b** Comparison of PS3 criterion application at any strength level (purple) and BS3 criterion application at any strength level (orange) to variants ultimately classified as variants of uncertain significance (VUS) in the pilot variant classification of each VCEP. **c** Comparison of the final classification (P, LP, VUS, LB, or B) of pilot variants with PS3/BS3 criteria (at any strength level). The *CDH1* VCEP and the Inherited Cardiomyopathy-*MYH7* VCEP did not use BS3 evidence in the interpretation of any pilot variants.

### CDH1 VCEP

The *CDH1* VCEP set guidelines for functional studies of the E-cadherin protein encoded by *CDH1* (Table 1; see also Additional File 1) [3]. Loss-of-function variants in the *CDH1* gene have been associated with hereditary diffuse gastric cancer [9] through a loss of cell adhesion and an increase in cell motility [10]. *In vitro* studies commonly test *CDH1* variants for retention of two main functions: cell-cell adhesion and invasion suppression, through aggregation assays or collagen invasion assays, respectively [11]. The *CDH1* VCEP evaluated 49 variants in their pilot study and assigned the PS3 criterion to four [3]. The only approved assays were those that measure abnormal splicing of the *CDH1* gene, as this measures one of the main disease mechanisms. For the purpose of this analysis, we only assessed functional studies that evaluate the effect of *CDH1* variants on protein function and not those assessing splicing variation (see Methods and Discussion).

This VCEP also reviewed literature studying the effect of missense variants and identified 14 variants with two or more published “abnormal” functional assay results, six of which were included in the pilot set. However, this VCEP ultimately decided these assays (aggregation/invasion, wound closure, and proximity ligation) were not sufficient predictors of pathogenicity, in part because none of the 14 variants were found in a large database of *CDH1* variants from individuals with disease [3]. To better understand why the VCEP deemed these assays poor predictors of pathogenicity for missense variants, we evaluated each functional assay the VCEP considered (Fig. 2; see also Additional File 2: Tables S1 and S2). We then compared the findings from these functional studies to assertions in ClinVar, both from the VCEP and other clinical labs (Additional File 2: Table S3), to examine if the functional assays that tested *CDH1* missense variants could predict pathogenicity. While most clinical lab entries in ClinVar did not specify which rule codes they used in their interpretation, many commented on the functional data. Only one of the 14 variants analyzed had a likely pathogenic assertion, while the remaining variants were classified as benign (5), VUS (8), conflicting (1), or not listed in ClinVar (3). We also noted that while each assay instance incorporated wild type and mock controls, no known pathogenic or benign controls were used to validate the assays. This limited validation coupled with an absence of identified definitively pathogenic missense variants makes it difficult to determine the positive predictive value of these assays and likely contributed to the VCEP not approving any existing functional studies of missense variants.

**Fig. 2.**
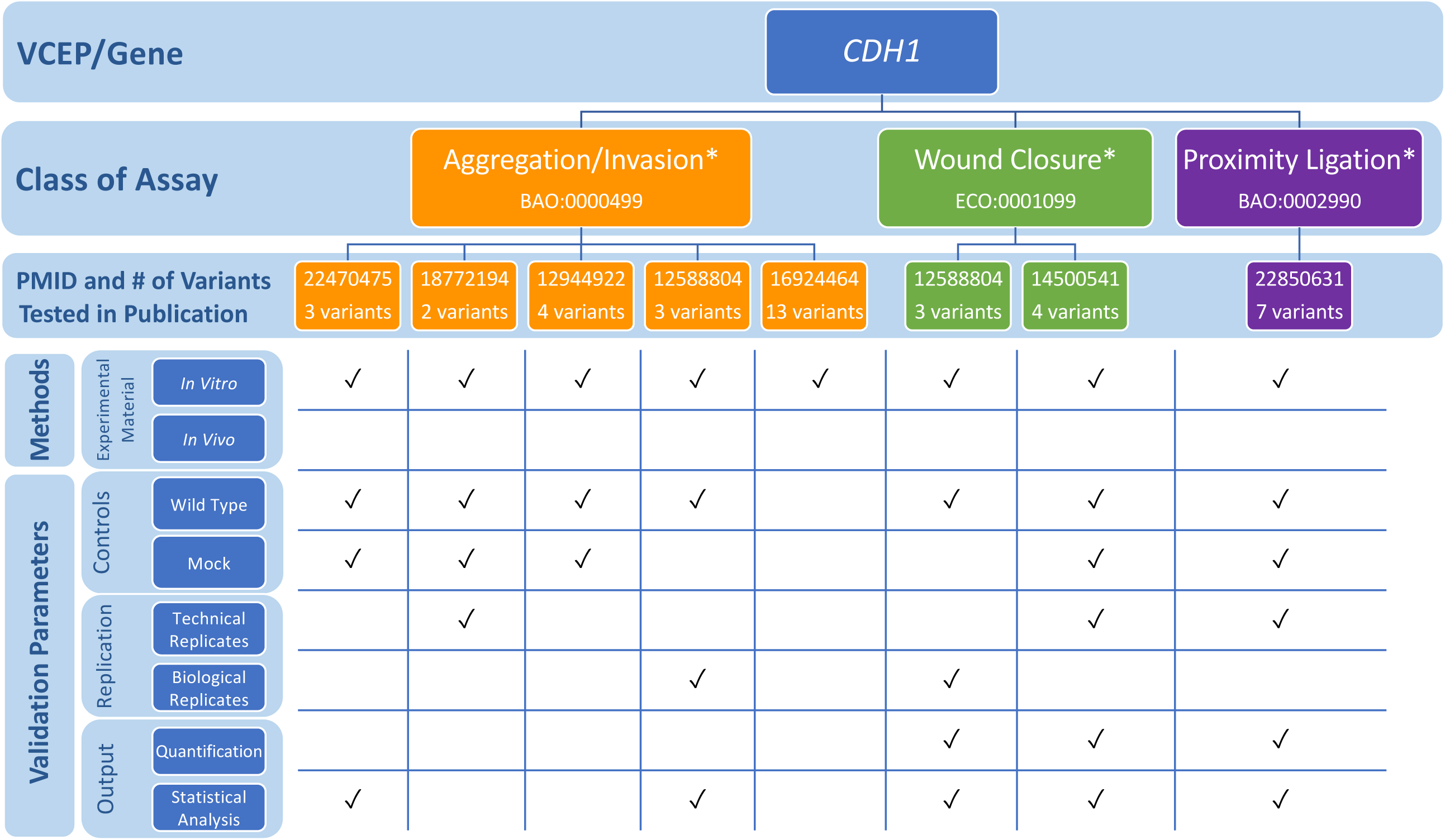
Representative findings from curation of specific instances of *CDH1* functional assays. We assessed the following methods and validation parameters of aggregation/invasion, wound closure, and proximity ligation assays: experimental material, controls, replication, and output. *Indicates assays not approved by the Variant Curation Expert Panel (VCEP). See Additional File 2: Tables S1 and S2 for full primary literature curation results.

### Hearing Loss VCEP

The Hearing Loss (HL) VCEP developed disease-specific recommendations for variant interpretation in the context of nine genes commonly associated with hearing loss: *CDH23, COCH, GJB2, KCNQ4, MYO6, MYO7A, SLC26A4, TECTA*, and *USH2A* (Table 1; see also Additional File 1) [5]. A single assay type, a variant-specific knock-in mouse model recapitulating the hearing loss phenotype, was the only functional study approved for PS3 application at the strong level. Additional guidelines for PS3/BS3 application at reduced strength levels were given for three genes: *COCH, GJB2*, and *SLC26A4*. Given the heterogeneity in disease mechanism underlying the multiple types of hearing loss, each gene was associated with a unique set of approved functional assays (Table 1). The HL VCEP calculated the positive and negative predictive value of functional assays commonly used to assess variants in these three genes (*COCH, GJB2, SLC26A4*) by comparing published assay results with ClinVar classifications [5]. For a P or LP ClinVar variant, an “abnormal” assay result compared to wild type was considered a true positive, while an assay result similar to wild type was considered a false negative. Similarly, for a variant classified as B or LB in ClinVar, a wild type-like assay result was considered a true negative, while an “abnormal” result was considered a false positive.

In the VCEP pilot variant classification of 41 variants, PS3 (at any strength level) was applied to eight variants and BS3 (at a supporting strength level) was applied to two variants. The VCEP did not cite any mouse models in their final variant curations, despite previous reports of mouse models generated for two pilot variants (*GJB2* c.109G>A and *SLC26A4* c.919-2A>G) [20,21]. Assays testing transport capability (electrical coupling, dye transfer, anion transport) were the most commonly used functional evidence (applied as PS3/BS3 assertions at reduced strength for eight variants). We assessed each of the 31 specific instances of these assays cited by the VCEP, in which some variants were evaluated more than once, to determine how often the parameters defined by the VCEP were satisfied (Fig. 3; see also Additional File 2: Tables S4 and S5). While all instances [22,23,32–41,24,42–48,25–31] tested a wild type control, water-injected or non-transfected controls were less consistently used (24/31), despite the VCEP’s stated requirement. Statistical testing was included in 17/31 cited instances of assay. In particular, no statistical analysis was done for the dye transfer assays, possibly because the results of this test are qualitative rather than quantitative. Finally, the HL VCEP applied PS3_supporting to a variant in an additional gene not given assay-specific recommendations, *KCNQ4* c.853G>A p.(Gly285Ser). Two instances of an electrical coupling assay [49,50] showing little to no electrical current in cells expressing *KCNQ4* p.Gly285Ser were used as evidence for PS3_supporting. Although no specific guidance was given for *KCNQ4* variant interpretation, functional assays with limited validation were generally approved by the VCEP at the PS3_supporting level for all hearing loss-associated genes.

**Fig. 3.**
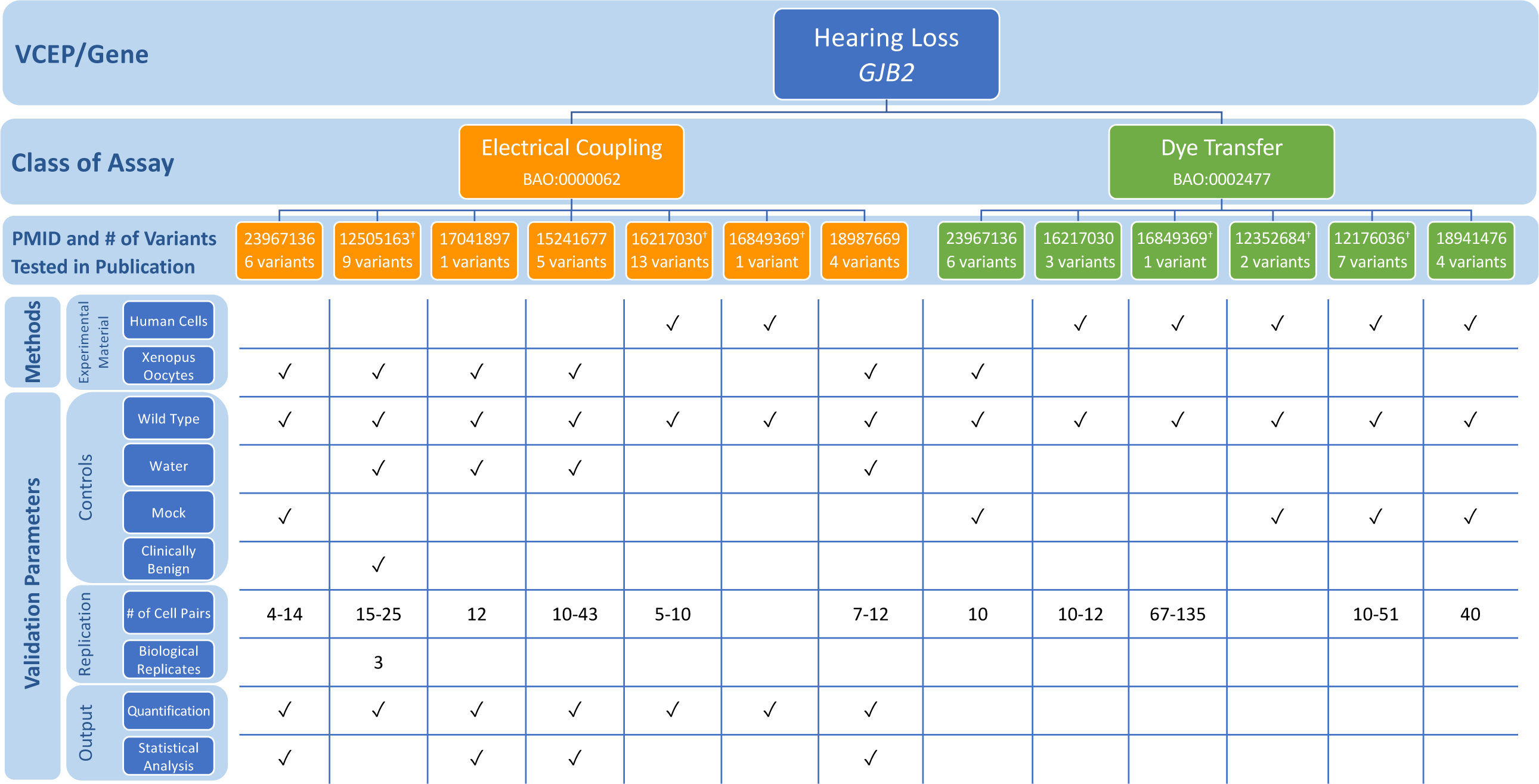
Representative findings from curation of specific instances of *GJB2* functional assays. We assessed the following methods and validation parameters of electrical coupling and dye transfer assays: experimental material, controls, replication, and output.^†^Indicates specific instance of assay cited by the Variant Curation Expert Panel (VCEP) as evidence for the PS3/BS3 criteria in pilot variant classification. See Additional File 2: Tables S4 and S5 for full primary literature curation results.

A variant in the pilot set, *SLC26A4* c.349C>T p.(Leu117Phe), was assigned BS3_supporting, but given an overall pathogenic classification (and, post-publication, downgraded to likely pathogenic in the ClinGen Evidence Repository). Although a radioactive anion isotope transport assay [46] found no statistically significant difference in rate of iodide efflux in cells expressing the *SLC26A4* variant, the VCEP reasoned that the functional assay may not assess all ion transport functions of the protein or fully reflect the biological environment, and noted that the benign functional evidence at a supporting level was not considered to be in conflict with other pathogenic evidence leading to the final classification. While the VCEP did not give specific recommendations for handling conflicting criteria, this case suggests that functional assays, even if VCEP-approved, are limited in their ability to test all functions of a protein. Functional evidence, especially evidence supporting BS3 criteria, must be weighed with other types of evidence in making an overall classification determination (see Discussion).

### Inherited Cardiomyopathy (MYH7) VCEP

The Inherited Cardiomyopathy VCEP published recommendations for interpretation of variants in *MYH7*, encoding α (alpha) cardiac myosin heavy chain, a gene associated with multiple forms of cardiomyopathy (dilated, hypertrophic, and restrictive) (Table 1; see also Additional File 1) [6]. The expert panel reviewed published functional evidence for their 60 pilot variants to determine which assays qualified for PS3/BS3 evidence. After evaluating *in vivo* and *in vitro* functional evidence for 23 of these variants, they approved only *in vivo* mammalian, variant-specific knock-in models to serve at the strong level and applied this evidence to four variants. Given the poor predictive value of the 16 *in vitro* assays evaluated in their review, no *in vitro* assays were approved at any strength level and were not cited as evidence for any pilot variants.

When assessing the various functional assays this expert panel reviewed but ultimately did not approve, we noted that the *MYH7* c.1208G>A p.(Arg403Gln) variant was tested in many of the functional studies. We used this variant to compare the characteristics of the assays that this VCEP did approve for use as evidence of PS3/BS3 (knock-in mouse model) to those that were not approved (*in vitro* motility assay and ATPase assay) (Fig. 4; see also Additional File 2: Tables S6 and S7). The first knock-in mouse model of hypertrophic cardiomyopathy introduced the c.1208G>A p.(Arg403Gln) variant into the endogenous murine *Myh7* [51]. The mice had a heart phenotype similar to hypertrophic cardiomyopathy that was recapitulated in multiple instances, which reported defective myocyte function and development of cardiac hypertrophy and lethal cardiomyopathy [36,52–57] in mice bearing an p.Arg403Gln *Myh7* variant. The VCEP considered this strong evidence for pathogenicity. We also reviewed two classes of *in vitro* functional assay commonly used to assess the effect of *MYH7* c.1208G>A p.(Arg403Gln), but not approved by the VCEP: the *in vitro* motility assay and the ATPase assay. The *in vitro* motility assay measures the velocity of actin filament sliding on a surface coated with myosin, a motion required for normal muscle contraction *in vivo* [58,59], while the ATPase assay measures the enzymatic function of ATP exchange required for force generation [60,61]. We examined several instances of each assay type [57,62–70] and noted heterogeneity in the source of myosin used, as well as a general lack of controls with known effect (other than wild type) for comparison to the variant myosin [57,62–70]. Furthermore, distinct instances of this assay examining the c.1208G>A p.(Arg403Gln) *MYH7* variant yielded conflicting results, with some studies finding increased actin filament velocity [57,65–69] or ATPase activity [57,66,67] and others reporting decreased actin filament velocity [62–64,70] or ATPase activity [64,68,70]. Poor reproducibility of the motility assay has been previously reported [71] and is thought to arise, at least partially, due to technical complications in myosin isolation. Ultimately, this case study demonstrates that poor reproducibility across instances of an assay class complicates the interpretation of the results in aggregate and no evidence from this assay class was approved for application.

**Fig. 4.**
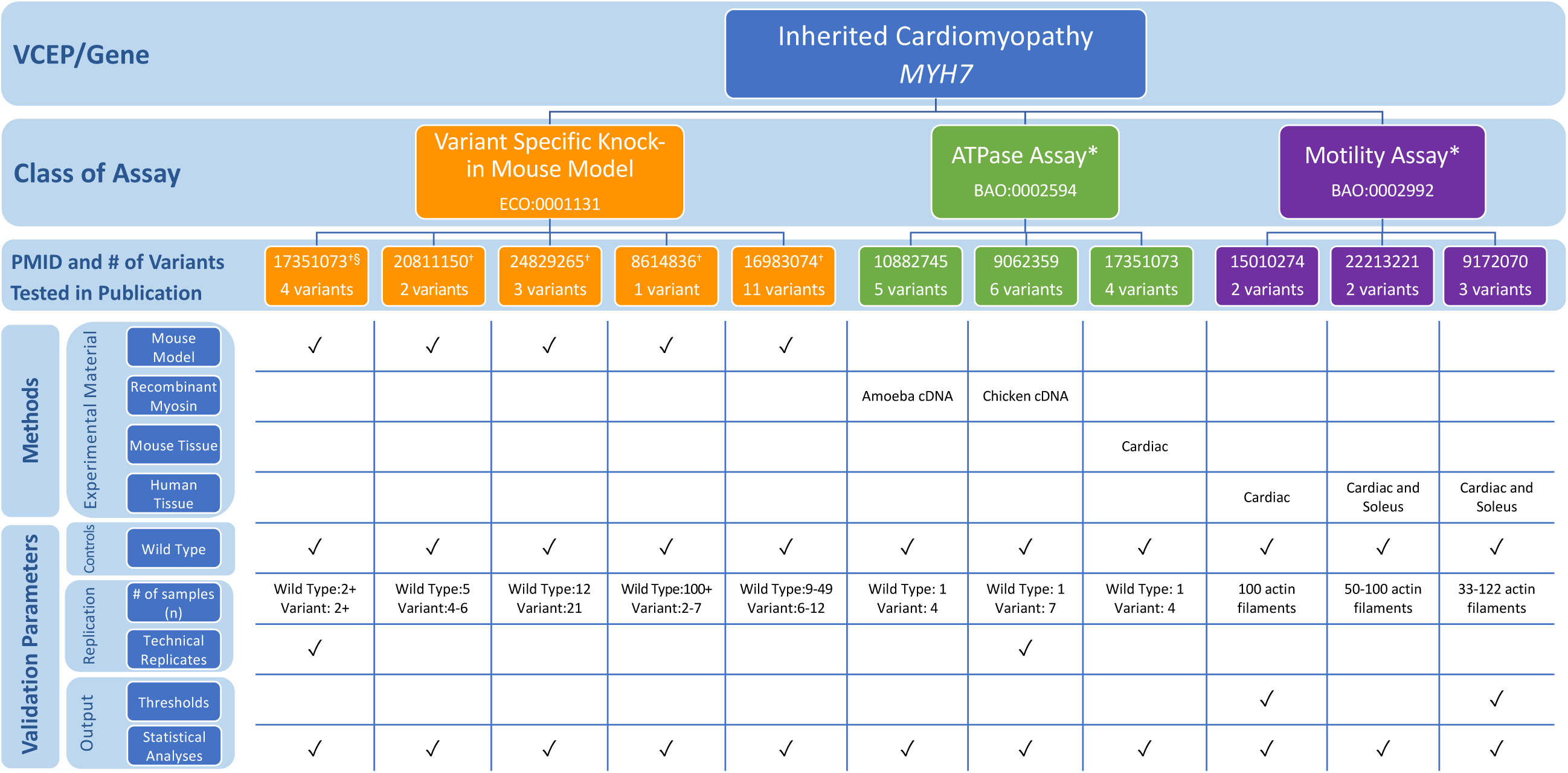
Representative findings from curation of specific instances of *MYH7* functional assays. We assessed the following methods and validation parameters of variant specific knock in mouse models, ATPase activity, and motility assays: experimental material, controls, replication, and output. *Indicates assays not approved by the Variant Curation Expert Panel (VCEP). ^†^Indicates specific instance of assay cited by the VCEP as evidence for the PS3/BS3 criteria in pilot variant classification. See Additional File 2: Tables S6 and S7 for primary literature curation results.

### PAH VCEP

The *PAH* VCEP published functional study guidelines for variants in the phenylalanine hydroxylase (*PAH*) gene associated with phenylketonuria (PKU) (Table 1; see also Additional File 1) [7]. They reviewed existing literature for functional studies and approved a well-established *in vitro* PAH enzyme activity assay involving expression of the variant allele in cultured cells and measurement of the variant enzyme activity in comparison to wild type activity. *In vitro* PAH enzyme activity correlates with the severity of the PKU phenotype [72]. A threshold of 0-50% residual enzyme activity compared to wild type was recommended for evidence of abnormal activity sufficient for PS3 application [7]. The VCEP assessed 85 variants in their pilot study and assigned PS3 to 29 variants with residual PAH activity values of ≤50% compared to wild type. The VCEP did not recommend the use of other assays described in the primary literature measuring *PAH* expression or protein folding, aggregation, or stability [73].

To assess use of the enzyme activity assay by the VCEP, we evaluated specific instances of assays measuring PAH activity cited as evidence in their pilot variant classification. We noted several discrepancies in assay methodology among the different research groups (Fig. 5; see also Additional File 2: Tables S8 and S9). In most instances, *PAH* variants were expressed in COS monkey kidney cells and enzyme activity was measured in cell extracts [73–79], though some expressed the variant in *E. coli* and measured enzyme activity of the purified protein [73,80,81]. In some instances, a synthetic cofactor 6-MPH4 [73,74,76,79,82] was used in place of the natural cofactor BH4 [76–78,80,81,83]. The method for measuring the conversion of phenylalanine to tyrosine also differed among experiments, with early researchers using paper chromatography or thin layer chromatography (TLC) and visualizing the results with autoradiography and quantifying via a liquid scintillation counter [73,75,79,83]. As technology advanced, experiments used high performance liquid chromatography (HPLC) with fluorometric detection [80,84] or the more sophisticated method of liquid chromatography measured with electrospray ionization tandem mass spectrometer [78].

**Fig. 5.**
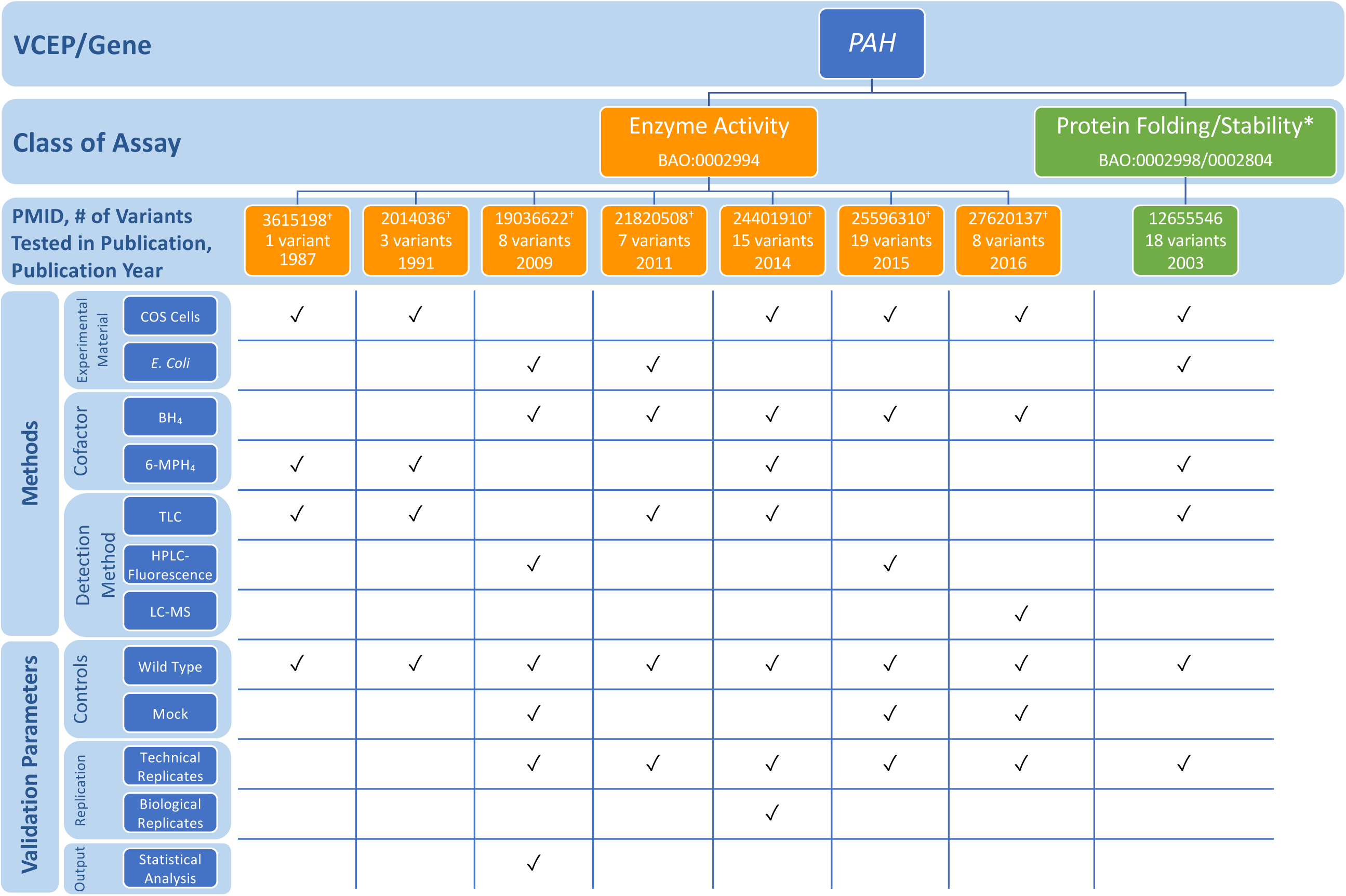
Representative findings from curation of specific instances of *PAH* functional assays. We assessed the following methods and validation parameters of enzyme activity and protein folding/stability assays: experimental material, cofactor, detection method, controls, replication, and output. *Indicates assays not approved by the Variant Curation Expert Panel (VCEP). ^†^Indicates specific instance of assay cited by the VCEP as evidence for the PS3/BS3 criteria in pilot variant classification. Abbreviations: TLC: Thin-layer chromatography; HPLC-Fluorescence: High-performance liquid chromatography coupled with fluorescence detection; LC-MS: Liquid chromatography coupled with mass spectrometry. See Additional File 2: Tables S8 and S9 for full primary literature curation results.

Given the differences in assay methodologies in instances of the PAH enzyme activity assay, we concluded that PAH activity results may vary among different instances of the assay analyzing the same variant. To test this, we compared enzyme activity results from multiple sources to the final enzyme activity cited by the VCEP as evidence for the interpretation of a given variant. One such source, a meta-analysis of *in vitro* PAH enzyme activity assays of 87 *PAH* variants from 49 publications also noted different methodologies for measuring PAH activity, including different cell expression systems, cofactors, temperatures, reaction times, and measurement methods, and variation in the final assay result [72]. Of the 29 variants assessed in the VCEP pilot study and ultimately assigned PS3, 18 had more than one result reported in the sources we reviewed (Table 3). We observed consistency in the activity levels of more severe variants that were often identified in individuals with classical PKU, but the milder variants had a wide range of reported enzyme activity levels. Of note, nine variants had at least one report of enzyme activity ≥50% of wild type, which would exceed the VCEP-established cut off and conflict with the VCEP-cited evidence in support of a PS3 assertion. The variation in enzyme activity levels may be dependent on experimental design, thus further guidance is needed on the most appropriate method to measure activity level and/or on how to resolve conflicting results.

**Table 3:** Comparison of PAH enzyme activities reported in a meta-analysis publication Himmelreich et al., 2018, the PAH locus-specific database (PAHvdb), and the *PAH* Variant Curation Expert Panel (VCEP) variant evidence for PS3 assertion.

| *PAH* Variant |  | % Enzyme Activity by Source |  |  |  |
| --- | --- | --- | --- | --- | --- |
| cDNA | Protein | Himmelreich et al., 2018<br>(PMID: 30037505) |  | *PAHvdb* | VCEP Evidence for<br>PS3 Assertion |
|  |  | Literature Review | Assay* |  |  |
| c.194T>C | p.(Ile65Thr) | 26, 27, 33, 48, **60**<sup>†</sup> |  | 33 | 25 |
| c.311C>A | p.(Ala104Asp) | **67**<sup>†</sup> | **77** | 27 | 26 |
| c.473G>A | p.(Arg158Gln) | 2<sup>†</sup>, 5, 9, 10, 29 |  | 10 | 0.2-1.8 |
| c.533A>G | p.(Glu178Gly) | 31 |  | 39 | 39 |
| c.721C>T | p.(Arg241Cys) | 25 | **57** | 25 | 25 |
| c.754C>T | p.(Arg252Trp) | 0, 0<sup>†</sup> | 15 | 0 | 1 |
| c.755G>A | p.(Arg252Gln) | 24 |  | 3 | 3 |
| c.782G>A | p.(Arg261Gln) | 27, 30, 43, 47, 52<sup>†</sup> | 23 | 44 | 15-30 |
| c.842C>T | p.(Pro281Leu) | 0, 1, 1<sup>†</sup> |  | 2 | 2 |
| c.898G>T | p.(Ala300Ser) | 32 | **65** | 31 | 31 |
| c.916A>G | p.(Ile306Val) | 12<sup>†</sup> | 25 | 39 | 18 |
| c.926C>T | p.(Ala309Val) | **70** | 12 | 42 | 30 |
| c.1162G>A | p.(Val388Met) | 15, 23<sup>†</sup>, 43 | 83 | 28 | 15 |
| c.1208C>T | p.(Ala403Val) | 32, **100** | 33 | 66 | 43 |
| c.1222C>T | p.(Arg408Trp) | 0, 1, 3, 5 | 2 | 2 | 1.3-1.85 |
| c.1223G>A | p.(Arg408Gln) | 9<sup>†</sup>, 33, **55**, **84** | 41 | 46 | 46 |
| c.1238G>C | p.(Arg413Pro) | 2 | 11 | 35 | <3 |
| c.1241A>G | p.(Tyr414Cys) | 28, 38<sup>†</sup>, 50, **80** |  | 57 | 50 |

\*Indicates results generated in a PAH enzyme activity assay conducted in Himmelreich et al., 2018 (PMID: 30037505). Other results were derived as part of literature review conducted in Himmelreich et al., 2018.
<sup>†</sup>Indicates enzyme activity result generated by PAH expression in *E. Coli*. All other results were generated by PAH expression in COS cells.
Bolded values indicate measured enzyme activity levels above the >50% enzyme activity range specified by the VCEP and in direct conflict with the VCEP-cited enzyme activity value in support of PS3 assertion.

### PTEN VCEP

The *PTEN* VCEP outlined specific recommendations for seven accepted general classes of functional assays testing the effect of variants in this gene associated with hereditary cancer (Table 1; see also Additional File 1) [8]. In the VCEP pilot variant classification of 36 *PTEN* variants, PS3 was applied to four variants, PS3_supporting was applied to one variant, and BS3_supporting was applied to one variant. Phosphatase activity was the most commonly used assay (three of four variants assigned PS3), with a single study [85] testing the ability of purified proteins to dephosphorylate PIP3 *in vitro* used to support the PS3 assertion for all three variants. Each of these variants displayed >90% reduction in phosphatase activity, well below the VCEP-approved threshold of ≥50% reduction in protein activity compared to wild type PTEN, and were replicated in three independent experiments, but the VCEP-specified catalytically inactive control was not included (Fig. 6; see also Additional File 2: Tables S10 and S11). Two variants were classified PS3 or BS3 based on splicing assays [86,87], which we did not evaluate (see Methods). The final variant in the pilot set was assigned PS3_supporting based on altered protein localization [88].

**Fig. 6.**
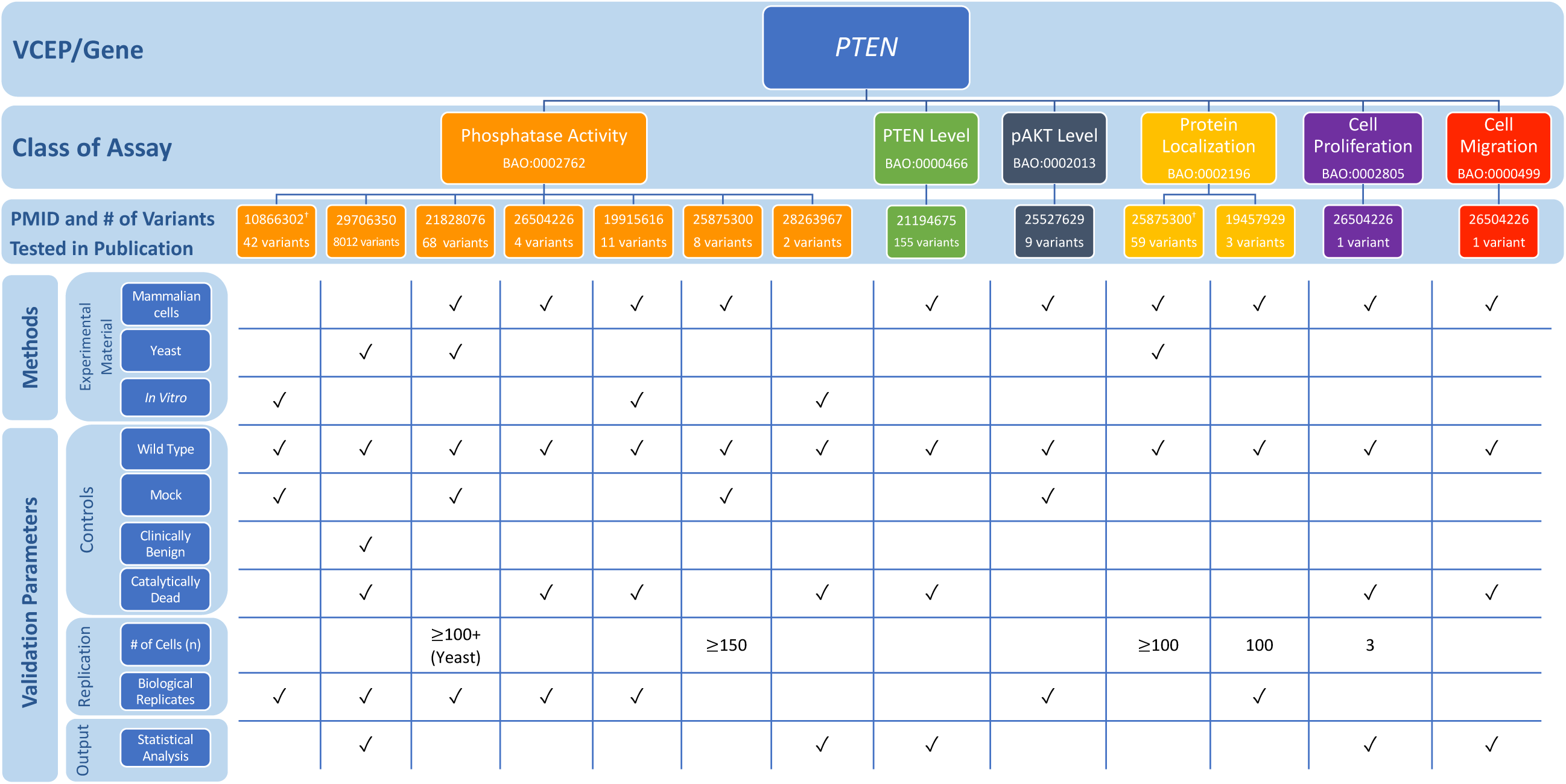
Representative findings from curation of specific instances of *PTEN* functional assays. We assessed the following methods and validation parameters of phosphatase activity, PTEN level, pAKT level, protein localization, cell proliferation, and cell migration assays: experimental material, controls, replication, and output. ^†^Indicates specific instance of assay cited by the Variant Curation Expert Panel (VCEP) as evidence for the PS3/BS3 criteria in pilot variant classification. See Additional File 2: Tables S10 and S11 for full primary literature curation results.

The VCEP cited a study that assessed PTEN protein levels in lymphoblast cell lines derived directly from individuals with Cowden syndrome [89]. Further guidance on the use of tissues and cell lines derived from affected individuals in functional assays is needed to inform application of this type of evidence (see Methods and Discussion). Additional guidance may also be necessary to interpret transgenic model organism evidence. Although the VCEP specified that this class of functional assay could be used in support of PS3_supporting, no studies of transgenic model organisms were used by the VCEP in pilot variant classification and the exact phenotypes required to use this type of evidence are unclear. Also of note was a high-throughput assay cited by the VCEP, but not used in pilot variant interpretation. This study used saturation mutagenesis to assess the pathogenicity of over 8000 *PTEN* variants, nearly all possible missense variants, by expressing *PTEN* in yeast cells and using cell growth rate as a readout for phosphatase activity [90]. The same three variants in the pilot set found to have reduced phosphatase activity in an *in vitro* phosphatase activity assay [85] were also tested in this high-throughput assay [90]. Fitness scores of all three of these variants were lower than that observed for wild type or “wild type-like” variants, suggesting agreement of this approach with small-scale *in vitro* assays.

### RASopathy VCEP

The RASopathy VCEP published recommendations for PS3/BS3 application in interpretation of variants in nine genes linked to RASopathy conditions: *BRAF, HRAS, KRAS, MAP2K1, MAP2K2, PTPN11, RAF1, SHOC2*, and *SOS1* (Table 1; see also Additional File 1) [9]. Assays measuring MAP2K1/2 and ERK1/2 phosphorylation [91,92,101–104,93–100] were the mostly commonly cited functional evidence in pilot variant classification (24/36 variants). The VCEP indicated that MAP2K1/2 and ERK1/2 activation should be measured both basally and following receptor tyrosine kinase stimulation, typically via epidermal or fibroblast growth factor addition (EGF and FGF). We noted disparities of assay instances with respect to whether measurements were taken in serum-starved cells, stimulated cells, or both, and in the method of stimulation (serum addition vs. purified EGF or FGF addition) (Fig. 7; see also Additional File 2: Tables S12 and S13). Direct quantification was not required, but was completed in many instances, as were statistical analyses.

**Fig. 7.**
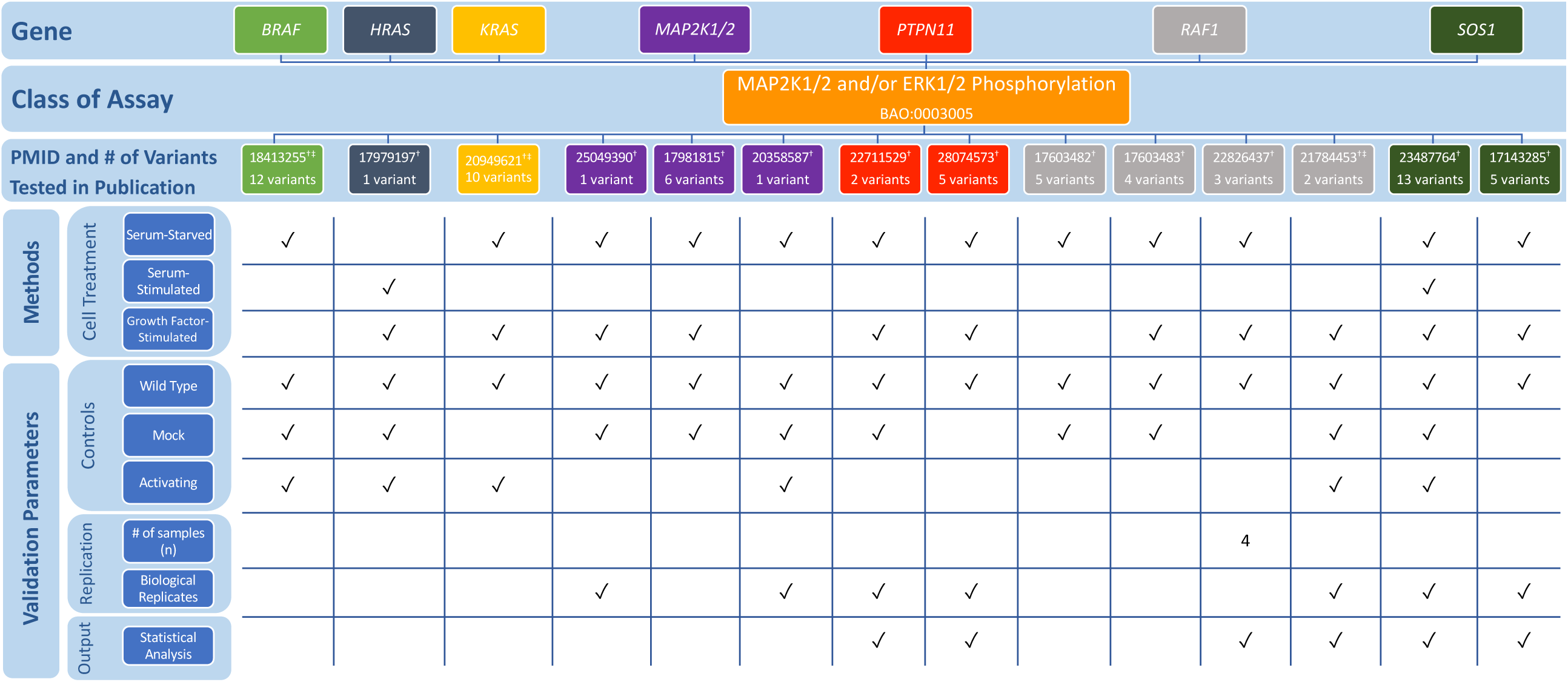
Representative findings from curation of specific instances of RASopathy functional assays. We assessed the following methods and validation parameters of MAP2K1/2 and ERK1/2 phosphorylation assays: cell treatment, controls, replication, and output.^†^Indicates specific instance of assay cited by the Variant Curation Expert Panel (VCEP) as evidence for the PS3/BS3 criteria in pilot variant classification. ^‡^Indicates specific instance of assay cited by the VCEP as evidence for the PS3/BS3 criteria in pilot variant classification for variants in multiple genes: PMID 18413255 cited as evidence for the PS3/BS3 criteria for variants in *BRAF* and *MAP2K1/2;* PMID 16439621 cited as evidence for PS3/BS3 for variants in *KRAS* and *MAP2K1/2;* PMID 21784453 cited as evidence for PS3/BS3 for variants in *RAF1* and *SOS1*. See Additional File 2: Tables S12 and S13 for full primary literature curation results.

Gain-of-function *BRAF* variants leading to an increase in kinase activity are most commonly associated with Cardio-facio-cutaneous syndrome; however, gain-of-function variants that result in reduced kinase activity and impaired stimulation of MAP2K1/2 and ERK1/2 phosphorylation have also been observed [101,105]. Although the VCEP specified that increased kinase activity could be used in support of PS3, no guidelines were given for variants that result in kinase impairment, despite their application of PS3 to variants with reduced BRAF kinase activity (e.g. *BRAF* c.1787G>T p.Gly596Val). Similarly, PS3 was applied to a *PTPN11* variant (c.1403C>T p.Thr468Met) with impaired phosphatase activity measured in different instances of the same assay type, but the VCEP only gave recommendations for variants that resulted in increased phosphatase activity. In two instances, the VCEP cited an *ELK* transactivation assay as evidence for PS3 application. While this assay was not explicitly approved by the VCEP, it appears to reflect the disease mechanism, as it measures the ability of *BRAF* to activate downstream transcription of *ELK* transcription factor.

## DISCUSSION

Our review of disease- and gene-specific functional assay evidence recommendations by six VCEPs highlighted a general uniformity across VCEPs in the approval of assays reflective of disease mechanism and, in some cases, the explicit exclusion of assays deemed to be poor predictors of variant pathogenicity. Our efforts also identified differences among VCEPs in parameter specification and evidence capturing, suggesting the need for a baseline guide for functional evidence assessment and a consistent criteria for capturing functional evidence. Along with the evidence curation criteria described in this study, standardized ClinGen criteria for functional assay evaluation should be developed to ensure consistency among VCEPs. Use of standard operating procedures for curating functional assay evidence could also improve transparency by encouraging complete recording of evidence used in variant classification, including documenting any conflicting evidence and whether a given piece of functional evidence for a variant was considered but not deemed appropriate, versus not evaluated at all.

We noted five recurring points of interest that will likely require further elucidation by the ClinGen Sequence Variant Interpretation Working Group to streamline functional evidence interpretation: (1) methodology for estimating the predictive power of assays, (2) consideration of splicing assays within PS3/BS3 criteria, (3) use of functional data from experimental materials derived from affected individuals, (4) unclear recommendations for creation and interpretation of model organism evidence, and (5) limited guidance for conflicting evidence.

Two VCEPs, Hearing Loss (HL) and *CDH1*, detailed their approach for estimating the predictive power of assays to determine which assays should be approved for use as PS3/BS3 evidence. The HL VCEP calculated the positive and negative predictive value of functional assays commonly used to assess variants in three genes (*COCH, GJB2, SLC26A4*) by comparing published assay results with ClinVar classifications [5], while the *CDH1* VCEP compared published assay results with data from affected individuals (see *CDH1* VCEP and Hearing Loss VCEP in Results) [4]. Importantly, these estimations were limited by the number of variants assessed, with 10 to 23 variants analyzed per hearing loss-related assay or by its reliance on previous identification of the variant in populations of affected individuals. Furthermore, the HL VCEP used aggregated results of multiple specific instances of a general class of assay, rather than assessing each instance and its validation parameters independently. In our view, the predictive value of a functional assay is most reliably determined using variants of known pathogenic or known benign interpretation (interpreted as such without using functional evidence) in the same instance of the assay, rather than attempting a *post hoc* calculation across different instances of the same assay. Clearly, additional guidance on appropriate methods for estimating the predictive power of assays is needed.

For the purpose of this analysis, we defined functional assays as systematic experiments (either *in vitro* or *in vivo*) used to elucidate the function of a protein in a cellular pathway or biological process [106]. With this in mind, we did not curate splicing assay evidence, despite splicing assessment being explicitly approved by the *CDH1, PAH*, and *PTEN* VCEPs and implicitly approved by the HL VCEP (via use of splicing evidence in interpretation of a pilot variant). While these assays can provide evidence of abnormal splicing and confirm results from *in silico* predictors, it does not directly test the function of the protein and, as a result, we suggest that splicing evidence represents a distinct type of evidence that may require separate interpretation recommendations.

We also observed relatively frequent citation of functional studies using cells or tissue derived from affected individuals in the primary literature used as evidence for PS3/BS3 criteria. As reasoned by Strande et al. [107], studies conducted using tissue or cells from an affected individual can provide high level information about the clinical phenotype (biochemical or enzymatic dysfunction), but not the variant-level effect, as the variant being tested cannot be isolated from other variants present in the individual’s genome. In general, this evidence may be better suited as evidence for the application of PP4 (supporting evidence of variant pathogenicity based on the individual’s specific phenotype as it relates to a disease).

Knock-in animal models were approved in some capacity by four of the six VCEPs; however, we noted a lack of guidance in their recommendations for model creation and interpretation. Some VCEPs gave no specifications for the number or type of different strains that should be used, the number of individual organisms that should be analyzed, or the features the animal must display to sufficiently recapitulate the disease phenotype. It was also unclear if studies using cells or tissue derived from a model organism for *in vitro* experiments should be considered model organism evidence. For example, although the Inherited Cardiomyopathy VCEP only approved mammalian variant-specific knock-in models, we noted at least one study used as pilot variant classification evidence that created a mouse model but did not directly assess the phenotype of the organism [69]. Instead, this study used myosin derived from these mice for *in vitro* assays of myosin motility and ATPase activity. Additional guidance on the interpretation of model organisms is necessary to reduce variability in evidence interpretation and application.

Our primary literature curation uncovered multiple instances of conflicting functional evidence for a single variant, yet only the HL VCEP provided guidance on interpretation of conflicting evidence from functional studies, suggesting “no criteria should be applied if multiple assay results do not agree” [5]. A striking example of the prevalence of conflicting data can be seen in different instances of the *PAH* VCEP-approved enzyme activity assay (Table 3). Variation in study design likely contributed to the wide range of activity levels observed for the same variant; however, this was not addressed by the VCEP and it is unclear how the VCEP selected which activity level to ultimately use as evidence for the PS3 criteria. This highlights the importance of not solely approving all assays of a given class, but rather evaluating the specific result of an assay in the context of that assay’s validation. In addition to conflicts between functional evidence, we also noted a need for guidance surrounding cases of functional evidence that conflict with other types of evidence gathered for a variant. The ACMG/AMP guidelines suggest that a variant with conflicting evidence should be classified as a VUS [1]. A Bayesian reinterpretation of ACMG/AMP guidelines specified a method for weighting combinations of pathogenic and benign evidence [108] that may help to solve this problem. For example, the Hearing Loss VCEP classified the *SLC26A4* variant c.349C>T as pathogenic (and later downgraded the variant to likely pathogenic), despite also applying BS3_supporting criteria to the variant. In doing so, the VCEP acknowledged that the anion isotope transport assay cited as evidence may not assess all aspects of protein function. Although not explicity stated, we infer that the VCEP did not want a “supporting” piece of evidence to call into question the overall interpretation, when other lines of evidence more strongly suggested a pathogenic interpretation. This example also raises the question of how functional assays that examine only one aspect of protein function should be interpreted when a variant demonstrates a normal result, and whether some form of combinatorial evidence from more than one class of assay should be required to support a benign interpretation.

Finally, through our curation of primary literature cited by the VCEPs, we observed that studies used as evidence for PS3/BS3 often did not satisfy all of the VCEP-recommended parameters (Fig. 2-7). Understandably, many functional assays have been performed in basic science laboratories for the purpose of understanding the gene, and not to provide clinically validated evidence of a pathogenic or benign classification. That being said, it is critical for VCEPs and others evaluating variants to approach this data critically and conservatively. While we suggest the development and implementation of criteria that sets baseline quality requirements, we also believe this finding demonstrates a need for the ACMG/AMP guidelines and VCEP recommendations surrounding PS3/BS3 criteria to be conveyed to research laboratories for incorporation into the study design of future research assays. Inclusion of pathogenic and benign controls, assay replication, and statistical analyses, among other practices, have the power to improve the clinical utility of studies conducted in research labs by aiding in clinical variant interpretation.

## CONCLUSIONS

In summary, our comparative analysis identified both commonalties and discrepancies among the functional assay evidence evaluation recommendations made by six ClinGen VCEPs. We observed multiples areas of discordance that warrant additional guidance, including setting a standard for basic validation parameters that should be fulfilled by functional studies, establishing if assays using experimental material derived from affected individuals are appropriate for PS3/BS3 evidence, and determining how conflicting evidence should be assessed. Although VCEP recommendations are an indispensable tool for the interpretation of functional evidence in a given disease area, further general guidance for functional evidence use is needed to take full advantage of the power of functional studies.

## Data Availability

All data generated or analyzed during this study supporting the conclusions of the article are included in this published article and its supplementary information files.

## LIST OF ABBREVIATIONS

ACMG: American College of Medical Genetics and Genomics
AMP: Association for Molecular Pathology
ATP: Adenosine Triphosphate
BH_4_: Tetrahydrobiopterin
DFNA9: autosomal dominant nonsyndromic deafness 9
DFNB1: autosomal recessive nonsyndromic deafness 1
DFNB3: autosomal dominant nonsyndromic deafness
DFNB4: autosomal recessive nonsyndromic deafness 4
BAO: Bioassay Ontology
B: benign
ClinGen: Clinical Genome Resource
ECO: Evidence and Conclusion Ontology
EGF: Epidermal Fibroblast Growth
FGF: Fibroblast Growth Factor
GO: Gene Ontology
HL: Hearing Loss
HPLC: high-performance liquid chromatography
LB: likely benign
LP: likely pathogenic
MONDO: Monarch Disease Ontology identifier
P: pathogenic
pAKT: phosphorylated AKT
PKU: Phenylketonuria
PMID: PubMed identifier
TLC: thin-layer chromatography
VCEP: Variant Curation Expert Panel
VUS: variant of uncertain significance

## DECLARATIONS

### Ethics approval and consent to participate

Not applicable

### Consent for publication

Not applicable

### Authors’ contributions

DMK, MKJ, SEB, CB, BCP, and JSB designed the study. DMK, SMM, and MKJ conducted the VCEP specification evaluation, variant identification, literature search, and curations. DMK, MKJ, SMM, SEB, CB, BCP, and JSB participated in data analysis. DMK, SMM, MKJ, and JSB wrote the original manuscript draft. DMK, MKJ, SMM, SEB, CB, BCP, and JSB reviewed and edited the final manuscript draft. All authors read and approved the final manuscript.

## Acknowledgements

We are grateful to ClinGen Sequence Variant Interpretation Working Group Chairs Leslie G. Biesecker (NIH/NHGRI) and Steven M. Harrison (Broad Institute) for their critical reading of this manuscript and valuable comments. We also appreciate helpful discussion with the ClinGen Functional Studies Team at Stanford University (Matt W. Wright, Michael A. Iacocca, Arturo Lopez Pineda, and Hannah Wand).

